# Hypothyroidism does not lead to worse prognosis in COVID-19: findings from the Brazilian COVID-19 registry

**DOI:** 10.1101/2021.11.03.21265685

**Authors:** Daniella Nunes Pereira, Leticia Ferreira Gontijo Silveira, Milena Maria Moreira Guimarães, Carísi Anne Polanczyk, Aline Gabrielle Sousa Nunes, André Soares de Moura Costa, Barbara Lopes Farace, Christiane Corrêa Rodrigues Cimini, Cíntia Alcantara de Carvalho, Daniela Ponce, Eliane Würdig Roesch, Euler Roberto Fernandes Manenti, Fernanda Barbosa Lucas, Fernanda d’Athayde Rodrigues, Fernando Anschau, Fernando Graça Aranha, Frederico Bartolazzi, Giovanna Grunewald Vietta, Guilherme Fagundes Nascimento, Helena Duani, Heloisa Reniers Vianna, Henrique Cerqueira Guimarães, Jamille Hemétrio Salles Martins Costa, Joanna d’Arc Lyra Batista, Joice Coutinho de Alvarenga, José Miguel Chatkin, Júlia Drumond Parreiras de Morais, Juliana Machado-Rugolo, Karen Brasil Ruschel, Lílian Santos Pinheiro, Luanna Silva Monteiro Menezes, Luciana Siuves Ferreira Couto, Luciane Kopittke, Luís César de Castro, Luiz Antônio Nasi, Máderson Alvares de Souza Cabral, Maiara Anschau Floriani, Maíra Dias Souza, Marcelo Carneiro, Maria Aparecida Camargos Bicalho, Mariana Frizzo de Godoy, Matheus Carvalho Alves Nogueira, Milton Henriques Guimarães Júnior, Natália da Cunha Severino Sampaio, Neimy Ramos de Oliveira, Pedro Ledic Assaf, Renan Goulart Finger, Roberta Xavier Campos, Rochele Mosmann Menezes, Saionara Cristina Francisco, Samuel Penchel Alvarenga, Silvana Mangeon Mereilles Guimarães, Silvia Ferreira Araújo, Talita Fischer Oliveira, Thulio Henrique Oliveira Diniz, Yuri Carlotto Ramires, Evelin Paola de Almeida Cenci, Thainara Conceição de Oliveira, Alexandre Vargas Schwarzbold, Patricia Klarmann Ziegelmann, Roberta Pozza, Magda Carvalho Pires, Milena Soriano Marcolino

## Abstract

**Background:** It is not clear whether previous thyroid diseases influence the course and outcomes of COVID-19. The study aims to compare clinical characteristics and outcomes of COVID-19 patients with and without hypothyroidism.

**Methods:** The study is a part of a multicentric cohort of patients with confirmed COVID-19 diagnosis, including data collected from 37 hospitals. Matching for age, sex, number of comorbidities and hospital was performed to select the patients without hypothyroidism for the paired analysis.

**Results:** From 7,762 COVID-19 patients, 526 had previously diagnosed hypothyroidism (50%) and 526 were selected as matched controls. The median age was 70 (interquartile range 59.0-80.0) years-old and 68.3% were females. The prevalence of underlying comorbidities were similar between groups, except for coronary and chronic kidney diseases, that had a higher prevalence in the hypothyroidism group (9.7% vs. 5.7%, p=0.015 and 9.9% vs. 4.8%, p=0.001, respectively). At hospital presentation, patients with hypothyroidism had a lower frequency of respiratory rate > 24 breaths per minute (36.1% vs 42.0%; p=0.050) and need of mechanical ventilation (4.0% vs 7.4%; p=0.016). D-dimer levels were slightly lower in hypothyroid patients (2.3 times higher than the reference value vs 2.9 times higher; p=0.037). In-hospital management was similar between groups, but hospital length-of-stay (8 vs 9 days; p=0.029) and mechanical ventilation requirement (25.4% vs. 33.1%; p=0.006) were lower for patients with hypothyroidism. There was a trend of lower in-hospital mortality in patients with hypothyroidism (22.1% vs. 27.0%; p=0.062).

**Conclusion:** In this large Brazilian COVID-19 Registry, patients with hypothyroidism had a lower requirement of mechanical ventilation, and showed a trend of lower in-hospital mortality. Therefore, hypothyroidism does not seem to be associated with a worse prognosis, and should not be considered among the comorbidities that indicate a risk factor for COVID-19 severity.

## Background

A global health crisis was established with the emergence of the coronavirus disease 19 (COVID-19).^1^ It is well known that individuals with some underlying medical conditions, such as cardiovascular diseases, cancer, obesity, diabetes and hypertension, are more likely to develop severe COVID-19, require hospitalization and intensive care, and have higher mortality rates.^2,3,4^ Nevertheless, other conditions such as asthma and Chagas disease surprisingly have not been related to more severe cases of COVID-19.^5^ However, many factors that interfere on COVID-19 clinical characteristics, symptoms and prognosis are still unknown and require further investigation to improve disease prevention and patient management.

It is not clear whether previous thyroid disease influences COVID-19 course.^6^ Previous studies suggested that patients’ thyroid status might have a direct impact on the course of COVID-19 due to the effects of thyroid hormone on multiple organs systems, including the cardiovascular and respiratory systems.^7^ Some studies have indeed observed that hypothyroidism may be associated with an increased risk for COVID-19 or a poorer prognosis in patients with the disease.^8,9^ However, these studies are limited due to small sample sizes and lack of comparison with patients without hypothyroidism.

Other studies have found that previous diagnosis of hypothyroidism or other thyroid diseases were not a risk factor for COVID-19 infection and were not associated with poorer outcomes of the disease.^10,11,12,13^ It is well known that critically ill patients with no previous history of thyroid dysfunction usually present alterations in thyroid hormones levels, a situation known as non-thyroidal illness syndrome (NTIS).^14^ Patients with NTIS typically present with decreased concentrations of plasma triiodothyronine (T3), low thyroxine (T4), and usually slightly decreased thyroid-stimulating hormone (TSH), which may be variable depending on the phase of the disease.^14^ In intensive care units (ICU), patients with conditions such as sepsis, major trauma or burn, and cardiac surgery, among others, NTIS has been associated with disease severity and mortality, without cause-effect association. Rather, NTIS is more likely to be seen as an adaptive mechanism to critical illnesses.^10,14,15^ As we could expect, NTIS has been frequently observed in COVID-19 patients, with the same pattern as in other critical conditions.^16,17,18^

Literature regarding the association between COVID-19 and previous hypothyroidism is still controversial, results may be biased by the inclusion of NTIS patients, and more studies with larger sample sizes and comparison with non-hypothyroidism patients are still needed. This study aims to compare the evolution of COVID-19 in patients with previous diagnosis of underlying hypothyroidism to patients without the disease, in order to understand if hypothyroidism is an independent risk-factor in the course of COVID-19.

## METHODS

### Study design and subjects

This study is a part of the multicentric cohort study Brazilian Covid-19 Registry, which included data from consecutive patients with confirmed COVID-19 diagnosis by RT-PCR or serological tests (IgM) from 37 hospitals.^19^ Patients were admitted from March 1st to September 31st, 2020.

For the present analysis, “cases” were patients with a clinical history of previously diagnosed hypothyroidism, on levothyroxine replacement therapy. From 7,762 patients with confirmed COVID-19 diagnosis, 555 (7.15%) had underlying hypothyroidism. Of those, 21 were excluded due to hospital admission for other reasons (not COVID-19), 3 were excluded due to pregnancy and 5 were transferred to other hospitals, with 526 hypothyroid patients remaining eligible for the study (Figure 1). Those patients were admitted in 31 hospitals, located in 15 cities (Belo Horizonte, Betim, Curvelo, Ipatinga, Teófilo Otoni, Divinópolis, Porto Alegre, Bento Gonçalves, Santa Cruz do Sul, Canoas, Santa Maria, Lajeado, Chapecó, Florianópolis and Botucatu) in 4 Brazilian states (Minas Gerais, Rio Grande do Sul, Santa Catarina and São Paulo). Of those, 13 were public, 15 private hospitals and 3 mixed hospitals. In order to compare patient outcomes and disease course, a paired analysis was made with matched controls with no previous history of hypothyroidism.

**Figure 1-.**
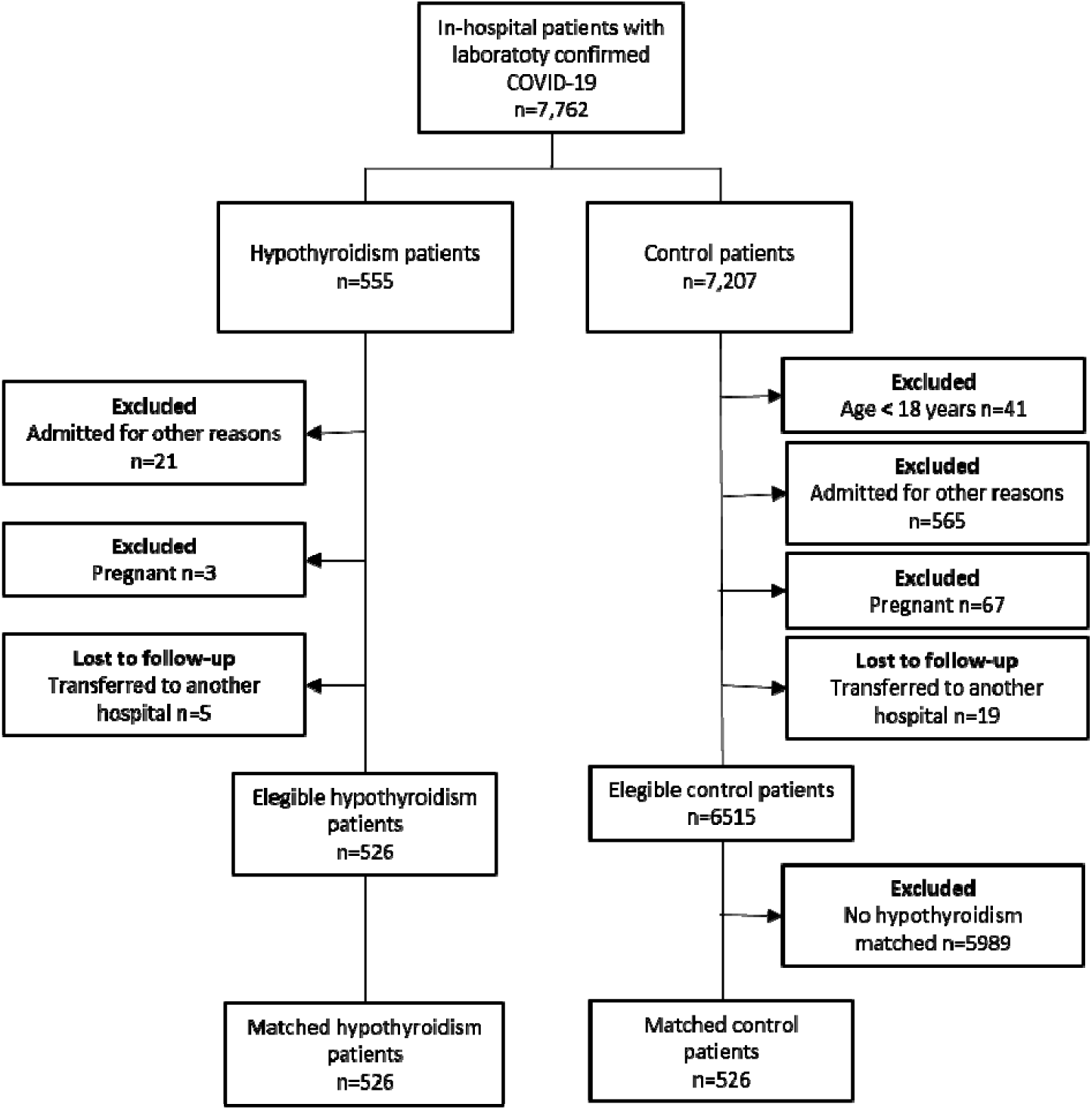
Flowchart of patients included in the study.

This study adheres to the STROBE guidelines (*Strengthening the Reporting of Observational Studies in Epidemiology*^20^), and it has been conducted by a protocol approved by the National Commission for Research Ethics^21^ (CAAE 30350820.5.1001.0008). Individual informed consent was waived due to the pandemic and to the fact that all data collected was unidentified, gathered through medical records.

### Data collection

Data was collected through the analysis of medical records by trained health professionals or undergraduate students (from Medical and Nursing schools) using Research Data Capture (REDCap) tools.^22^ In the medical records, data collected concerned demographic and clinical characteristics, comorbidities, medication in use prior to admission, COVID-19 symptoms, clinical evaluation at admission, laboratory and radiological exams, medication used during hospitalization and outcomes. Study protocol and definitions were published elsewhere.^23^

### Statistical analysis

Statistical analysis was performed using R software (version 4.0.2). To adjust for potential confounding variables, patients who had underlying hypothyroidism were matched to patients who had not underlying hypothyroidism (controls) on the basis of propensity score. Propensity score model was estimated by logistic regression, and included gender, age, number of comorbidities (hypertension, diabetes mellitus, obesity, coronary artery disease, heart failure, atrial fibrillation or flutter, cirrhosis, chronic obstructive pulmonary disease, cancer and previous stroke)^4^ and hospital. Patients from the control group were searched to find those who had the closest propensity score from the hypothyroidism group (within 0.17 standard deviations of the logit of the propensity score, on a scale from 0-1.00), using the MatchIt package in R software.

Categorical data were presented as absolute frequency and proportions, and continuous variables were expressed as medians and interquartile ranges. The Chi-Square and Fisher Exact tests were used to compare the distribution of categorical variables, and the Wilcoxon-Mann–Whitney test for continuous variables. Results were considered statistically significant if the two-tailed P-value was < 0.05.

## RESULTS

The study sample comprehended 526 individuals with previous hypothyroidism history and 526 matched controls. The median age was 70 yrs (59-80) and 68.3% were female. Most patients (85.8%) had at least one comorbidity, with the most common being hypertension (72.0%), diabetes mellitus (38.8%), obesity (18.9%) and heart failure (11,2%). In comparison to the controls, patients with underlying hypothyroidism had similar prevalence of comorbidities and unhealthy habits, except for a higher prevalence of coronary disease (9.7% vs. 5.7%, p=0.015) and chronic kidney disease (9.9% vs. 4.8%, p= 0.001) among the hypothyroidism group (Table 1).

**Table 1-.** Demographics and clinical characteristics of the study cohort.

|  | Hypothyroidism<br>N = 526<sup>1</sup> | Control patients<br>N = 526<sup>1</sup> | p-value<sup>2</sup> |
| --- | --- | --- | --- |
| **Age** (years) | 71.0 (60.0, 80.0) | 70.0 (59.0, 80.0) | 0.659 |
| **Female sex** | 356 (67.7%) | 363 (69.0%) | 0.643 |
| *Cardiovascular diseases* |  |  |  |
| Hypertension | 370 (70.3%) | 387 (73.6%) | 0.243 |
| Heart failure | 65 (12.4%) | 53 (10.1%) | 0.241 |
| Coronary artery disease | 51 (9.7%) | 30 (5.7%) | 0.015 |
| Atrial fibrillation/flutter | 36 (6.8%) | 37 (7.0%) | 0.903 |
| Stroke | 28 (5.3%) | 34 (6.5%) | 0.432 |
| *Respiratory diseases* |  |  |  |
| COPD | 42 (8.0%) | 44 (8.4%) | 0.822 |
| Asthma | 35 (6.7%) | 34 (6.5%) | 0.901 |
| *Metabolic diseases* |  |  |  |
| Diabetes mellitus | 215 (40.9%) | 193 (36.7%) | 0.164 |
| Obesity (BMI > 30kg/m2) | 94 (17.9%) | 95 (18.1%) | 0.936 |
| *Other conditions* |  |  |  |
| Chronic kidney disease | 52 (9.9%) | 25 (4.8%) | 0.001 |
| Cancer | 29 (5.5%) | 38 (7.2%) | 0.256 |
| Dementia | 16 (3.0%) | 11 (2.1%) | 0.330 |
| Rheumatological disease | 16 (3.0%) | 9 (1.7%) | 0.157 |
| Cirrhosis | 4 (0.8%) | 7 (1.3%) | 0.363 |
| Transplant | 3 (0.6%) | 3 (0.6%) | >0.999 |
| HIV infection | 2 (0.4%) | 4 (0.8%) | 0.687 |
| **Comorbidities** (total number) |  |  | 0.549 |
| 0 | 71 (13.5%) | 78 (14.8%) |  |
| 1 | 157 (29.8%) | 143 (27.2%) |  |
| 2 | 163 (31.0%) | 173 (32.9%) |  |
| 3 | 87 (16.5%) | 95 (18.1%) |  |
| 4 | 36 (6.8%) | 31 (5.9%) |  |
| >= 5 | 12 (2.3%) | 6 (1.2%) |  |
| **Toxic habits** |  |  |  |
| Alcoholism | 10 (1.9%) | 10 (1.9%) | >0.999 |
| Smoking | 100 (19.0%) | 115 (21.9%) | 0.251 |

<sup>1</sup>n (%); Median (IQR) <sup>2</sup>Pearson's Chi-squared test; Wilcoxon rank sum test; Fisher's exact test. COPD - Chronic obstructive pulmonary disease

The median duration of symptoms was 6 days (3-9 days) for both groups. The most predominant symptoms in disease presentation were dyspnea, fever and dry cough, being similar in both groups. Upon hospital presentation, patients with hypothyroidism had a lower frequency of respiratory rate over 24 breaths per minute (36.1% vs 42.0%; p=0.050), as well as lower proportion of mechanical ventilation (4.0% vs 7.4%; p=0.016) and of inotropics (24.3% vs 29.8%; p=0.044) requirement. The median peripheral oxygen saturation/fraction of inspired oxygen ratio (SF ratio) was slightly higher in patients with hypothyroidism than in the control group (428.6 vs. 423.8, p=0.034), as shown on Table 2.

**Table 2-.** Clinical characteristics at hospital admission.

|  | Hypothyroidism |  | Control patients |  | p-value<sup>2</sup> |
| --- | --- | --- | --- | --- | --- |
|  | N = 526<sup>1</sup> | Non missing cases | N = 526<sup>1</sup> | Non missing cases |  |
| Symptoms |  |  |  |  |  |
| Duration of symptoms (days) | 6.0 (3.0, 9.0) | 524 | 6.0 (3.0, 9.0) | 524 | 0.635 |
| Adynamic | 138 (26.2%) | 526 | 143 (27.2%) | 526 | 0.728 |
| Ageusia | 25 (4.8%) | 526 | 34 (6.5%) | 526 | 0.228 |
| Anosmia | 44 (8.4%) | 526 | 48 (9.1%) | 526 | 0.662 |
| Headache | 95 (18.1%) | 526 | 94 (17.9%) | 526 | 0.936 |
| Rhinorrhea | 72 (13.7%) | 526 | 79 (15.0%) | 526 | 0.538 |
| Diarrhea | 84 (16.0%) | 526 | 77 (14.6%) | 526 | 0.549 |
| Dyspnea | 298 (56.7%) | 526 | 316 (60.1%) | 526 | 0.260 |
| Fever | 278 (52.9%) | 526 | 247 (47.0%) | 526 | 0.056 |
| Hyporexia | 71 (13.5%) | 526 | 66 (12.5%) | 526 | 0.647 |
| Neurological manifestations | 14 (2.7%) | 526 | 21 (4.0%) | 526 | 0.229 |
| Myalgia | 147 (27.9%) | 526 | 130 (24.7%) | 526 | 0.234 |
| Nausea/vomiting | 83 (15.8%) | 526 | 69 (13.1%) | 526 | 0.220 |
| Productive cough | 71 (13.5%) | 526 | 79 (15.0%) | 526 | 0.481 |
| Dry cough | 262 (49.8%) | 526 | 268 (51.0%) | 526 | 0.711 |
| **Clinical assessment** |  |  |  |  |  |
| GCS < 15 | 84 (16.0%) | 526 | 83 (15.8%) | 526 | 0.933 |
| HR (bpm) | 85.0 (74.0, 96.0) | 505 | 87.0 (76.0, 98.0) | 507 | 0.169 |
| HR > 100 bpm | 116 (22.1%) | 526 | 131 (24.9%) | 526 | 0.275 |
| RR (bpm) | 20.0 (18.0, 23.0) | 443 | 20.0 (18.0, 24.0) | 430 | 0.216 |
| RR > 24 bpm | 190 (36.1%) | 526 | 221 (42.0%) | 526 | 0.050 |
| Sat O<sub>2</sub> (%) | 94.0 (91.0, 96.0) | 513 | 94.0 (91.0, 97.0) | 513 | 0.845 |
| Sat O<sub>2</sub> < 90% | 98 (18.6%) | 526 | 103 (19.6%) | 526 | 0.695 |
| Mechanical ventilation | 21 (4.0%) | 526 | 39 (7.4%) | 524 | 0.016 |
| Systolic blood pressure |  | 526 |  | 526 | 0.693 |
| SBP ≥ 90 mmHg | 482 (91.6%) |  | 464 (88.2%) |  |  |
| SBP < 90 mmHg | 25 (4.8%) |  | 28 (5.3%) |  |  |
| Inotropic use<sup>2</sup> | 19 (3.6%) |  | 34 (6.5%) |  |  |
| SF ratio<sup>2</sup> | 428.6 (339.3, 452.4) | 507 | 423.8 (304.7, 452.4) | 507 | 0.034 |

<sup>1</sup>Median (IQR); n (%), <sup>2</sup>Inotropic use at admission
SF ratio: peripheral capillary oxygen saturation/fraction of inspired oxygen (Spo<sub>2</sub>/Fio<sub>2</sub> Ratio)
GCS: Glasgow coma score; HR: heart rate; RR: respiratory rate; Sat O<sub>2</sub>: peripheral capillary oxygen saturation

Laboratory exams and biomarkers were similar between groups, except for D-dimer, which was lower in patients with hypothyroidism than the controls (Table 3).

**Table 3-.** Findings of laboratory exams.

|  | Hypothyroidism |  | Control patients |  |  |
| --- | --- | --- | --- | --- | --- |
|  | N = 526<sup>1</sup> | Non missing cases | N = 526<sup>1</sup> | Non missing cases | p-value<sup>2</sup> |
| Hemogram |  |  |  |  |  |
| Hemoglobin (g/L) | 12.6 (11.6, 13.9) | 515 | 12.9 (11.6, 13.8) | 509 | 0.301 |
| Leukocytes count (cels/mm<sup>3</sup>) | 6,785.0 (5,000.0, 9,302.5) | 512 | 6,940.5 (5,200.0, 9,132.5) | 510 | 0.317 |
| Neutrophiles (cels/mm<sup>3</sup>) | 4,737.0 (3,310.5, 7,155.8) | 498 | 5,204.0 (3,481.8, 7,281.2) | 488 | 0.205 |
| Lymphocytes (cels/mm<sup>3</sup>) | 1,100.0 (760.0, 1,503.5) | 499 | 1,041.0 (710.0, 1,421.0) | 489 | 0.136 |
| Platelet count (10<sup>9</sup>/L) | 195.0 (151.0, 244.8) | 508 | 196.3 (152.0, 261.0) | 505 | 0.499 |
| aPTT (secs/control) | 1.0 (0.9, 1.2) | 228 | 1.0 (0.9, 1.2) | 265 | 0.396 |
| Liver panel |  |  |  |  |  |
| Albumin (g/dL) | 3.4 (3.0, 3.7) | 90 | 3.3 (3.0, 3.6) | 104 | 0.283 |
| Total bilirubin (mg/dL) | 0.4 (0.3, 0.6) | 253 | 0.4 (0.3, 0.6) | 249 | 0.150 |
| Direct Bilirubin (mg/dL) | 0.2 (0.1, 0.3) | 251 | 0.2 (0.1, 0.3) | 249 | 0.013 |
| AST (U/L) | 37.4 (29.0, 55.0) | 326 | 38.0 (28.0, 52.0) | 334 | 0.750 |
| ALT (U/L) | 29.0 (20.0, 46.0) | 323 | 28.0 (19.0, 44.3) | 335 | 0.592 |
| Electrolytes |  |  |  |  |  |
| Calcium (mmol/L) | 1.1 (1.1, 1.2) | 138 | 1.1 (1.1, 1.2) | 167 | 0.298 |
| Potassium (mmol/L) | 4.2 (3.8, 4.6) | 476 | 4.1 (3.7, 4.5) | 485 | 0.065 |
| Sodium (mmol/L) | 137.0 (134.0, 139.7) | 474 | 137.7 (135.0, 140.0) | 482 | 0.020 |
| **Others** |  |  |  |  |  |
| BNP (pg/mL) | 78.0 (23.0, 230.5) | 47 | 110.0 (32.8, 312.0) | 48 | 0.218 |
| Creatinine (mg/dL) | 0.9 (0.8, 1.4) | 493 | 0.9 (0.7, 1.2) | 499 | 0.159 |
| CPK (U/L) | 81.5 (45.2, 145.8) | 150 | 74.5 (47.0, 189.9) | 149 | 0.668 |
| D-dimer* | 2.3 (1.2, 6.8) | 301 | 2.9 (1.2, 23.9) | 318 | 0.037 |
| Ferritin (ng/mL) | 80.1 (57.7, 142.0) | 19 | 66.0 (42.0, 175.3) | 17 | 0.447 |
| Fibrinogen (g/L) | 465.5 (334.5, 591.8) | 40 | 451.0 (343.0, 553.0) | 45 | 0.745 |
| Lactate dehydrogenase | 358.5 (255.2, 493.8) | 282 | 363.0 (270.0, 504.0) | 297 | 0.536 |
| NT-proBNP (pg/mL) | 310.0 (114.0, 785.7) | 55 | 302.0 (89.3, 1,197.5) | 70 | 0.895 |
| C reactive protein (mg/L) | 75.3 (32.1, 134.3) | 466 | 74.8 (34.3, 143.3) | 441 | 0.933 |
| PTTa (secs/control) | 1.0 (0.9, 1.2) | 228 | 1.0 (0.9, 1.2) | 265 | 0.396 |
| INR | 1.1 (1.0, 1.2) | 291 | 1.1 (1.0, 1.2) | 317 | 0.229 |
| Troponin* | 0.3 (0.1, 0.5) | 171 | 0.3 (0.1, 0.5) | 184 | 0.261 |
| Lactate | 1.4 (1.0, 1.9) | 311 | 1.4 (1.1, 1.9) | 321 | 0.427 |
| Urea (mg/dL) | 39.1 (28.0, 58.0) | 492 | 38.0 (28.0, 55.8) | 484 | 0.539 |

<sup>1</sup>Median (IQR); n (%) <sup>2</sup>Wilcoxon rank sum test; Fisher's exact test
aPTT :activated partial thromboplastin time; AST: aspartate transaminase; ALT: alanine aminotransferase; BNP B-type natriuretic peptide; CPK:creatine phosphokinase; NT-proBNP: N-terminal pro-brain natriuretic peptide; INR: international normalized ratio
\* Times the reference value.

As for medications used during hospitalization, hypothyroidism patients had a lower frequency of inotropes requirement than the control group (24.3% vs. 29.8%, p =0.044). The other medications were similar between groups (Table S1).

With regards to patient outcomes, the median hospital length of stay (8 vs 9 days; p=0.029) and mechanical ventilation requirement (25.4% vs. 33.1%; p=0.006) were lower in patients with hypothyroidism than in controls. A trend of lower in-hospital mortality was seen in patients with hypothyroidism (22.1% vs. 27.0%; p=0.062). The other outcomes were similar between groups, as shown in Table 4.

**Table 4-.** Patient outcomes.

|  | **Hypothyroidism**<br>**N = 526<sup>1</sup>** | **Control patients**<br>**N = 526<sup>1</sup>** | **p-value<sup>2</sup>** |
| --- | --- | --- | --- |
| Hospital length of stay, days | 8.0 (4.0, 14.0) | 9.0 (5.0, 14.2) | 0.029 |
| ICU, days | 197 (37.5%) | 224 (42.6%) | 0.089 |
| Time of ICU admission | 1.0 (0.0, 3.0) | 1.0 (0.0, 2.0) | 0.089 |
| Days at ICU | 8.0 (3.0, 15.0) | 8.0 (4.0, 17.0) | 0.293 |
| Mechanical ventilation, number of patients | 133 (25.4%) | 172 (33.1%) | 0.006 |
| Dialysis | 57 (10.8%) | 63 (12.0%) | 0.553 |
| Sepsis | 72 (13.7%) | 90 (17.1%) | 0.124 |
| Disseminated Intravascular coagulation | 0 (0.0%) | 3 (0.6%) | 0.249 |
| Decompensated chronic heart failure | 16 (3.0%) | 25 (4.8%) | 0.152 |
| Acute heart failure | 3 (0.6%) | 6 (1.1%) | 0.506 |
| Nosocomial infection | 48 (9.1%) | 48 (9.1%) | >0.999 |
| Miocardites | 0 (0.0%) | 3 (0.6%) | 0.249 |
| Hemorrhage | 10 (1.9%) | 9 (1.7%) | 0.817 |
| Vascular thrombosis | 18 (3.4%) | 22 (4.1%) | 0.519 |
| VTE | 16 (3.0%) | 21 (4.0%) | 0.403 |
| Arterial thrombosis | 2 (0.4%) | 1 (0.2%) | >0.999 |
| In hospital mortality | 116 (22.1%) | 142 (27.0%) | 0.062 |

<sup>1</sup>Median (IQR); n (%) <sup>2</sup>Wilcoxon rank sum test; Pearson's Chi-squared test; Fisher's exact test ICU (intensive care unit); VTE (venous thromboembolism)

## DISCUSSION

In this multicenter cohort of inhospital COVID-19 patients, those who had hypothyroidism had similar comorbidities, clinical manifestations and laboratory parameters when compared to the control group. Surprisingly, requirement for mechanical ventilation was remarkably lower in patients with underlying hypothyroidism, and there was a trend for lower hospital mortality when compared to controls, matched for age, sex, number of comorbidities and hospital.

Underlying individual comorbidities were similar between groups, except for chronic kidney disease and coronary artery disease. There has been a great interest in the relationship between thyroid dysfunction and kidney function in recent years.^24,25^ Thyroid hormones have shown to directly affect the kidneys, influencing renal growth and development, glomerular filtration rate, renal transport systems, as well as sodium and water homeostasis. An indirect effect through modifications in cardiac and vascular function and disruptions in the renin-angiotensin system is also believed to play an important role in reducing glomerular filtration rate through impaired renal autoregulation. At the same time, patients with autoimmune thyroid disorders are also at risk for immune-mediated glomerular diseases.^25^

As for the higher prevalence of coronary artery disease, the association between hypothyroidism and increased cardiovascular risk is controversial when thyroid-stimulating hormone (TSH) is normal.^26^ Nonetheless, it was demonstrated that patients with underlying hypothyroidism with a poor disease control have a worse lipidic profile and a higher prevalence of atherosclerosis, since thyroid hormones regulate lipid metabolism, especially TSH.^27,28,29^ At the same time, it is well established that a key-factor for adverse cardiovascular events prevention in patients with hypothyroidism is the achievement of symptom control, as well as levels of TSH and T4 within the reference values with the use of hormonal replacement therapy.^30^ Another hypothesis to explain the higher frequency of coronary artery disease is the higher prevalence of chronic kidney disease, which is associated with increased cardiovascular risk.

There was no difference in frequency and severity of COVID-19 symptoms between groups, including anosmia and ageusia. Despite the previous evidence of an effect of hypothyroidism at gustatory and olfactory perceptual pathways, including receptors, central olfactory and gustatory areas, and high order cognitive systems, previous cohorts studies did not observe a higher frequency of gustatory or olfactory COVID-19 symptoms in these patients. However, a case series has observed an independent association between hypothyroidism and a higher likelihood of persistent olfactory dysfunction among patients with COVID-19 (OR: 21.1; 95%[:CI: 2.0 to 219.4).^31^ Further studies are needed to investigate whether hypothyroidism is indeed associated with a higher risk of persistent symptoms.

Overall, laboratorial exams were similar between groups, except for D-dimer levels, which were slightly higher in the control group when compared to patients with hypothyroidism. However, the difference is not clinically relevant. These findings are different from a study from Wuhan, China, in the early phase of the pandemic, which observed that patients with thyroid dysfunction had persistently high levels of biomarkers for inflammatory response and cardiac injury. However, authors defined thyroid dysfunction by abnormal thyroid function tests results within three days from admission, and not previous underlying diseases.^8^

Current data shows that COVID-19 is associated with a broad spectrum of thyroid dysfunction, ranging from thyrotoxicosis to NTIS and hypothyroidism, that may worsen disease course and affect prognosis.^32,33,34,35,36^ This was observed even in mild to moderate COVID-19 cases, independently of SARS-CoV-2 viral load, age and inflammation and tissue injury markers.^37^ The cytokine storm and the dysregulated inflammation are believed to affect the thyroid gland, through the proinflammatory cytokines, which resembles the immune activation that occurs in immune-mediated thyroid diseases, causing damage and compromising function.^32^ It is also possible that thyroid abnormalities are caused by a direct effect of the virus, leading to subacute thyroiditis, a self-limiting destructive thyroiditis, or affecting angiotensin-converting enzyme 2 (ACE2) receptors thyroid follicular cells, leading to a thyroid malfunction.^32^ Other contributing factors may be the oxidative stress due to augmented reactive oxygen species generation, which are associated with NTIS, and changes in the intracellular redox state, that may disrupt deiodinase function by independent mechanisms, which might include depletion of the-as yet unidentified-endogenous thiol cofactor.^38^ Nevertheless, there is no evidence that the administration of thyroid hormones improves prognosis in those patients. We might also wonder that, in some studies that observed a correlation between thyroid dysfunction and more severe COVID-19 presentation, the alterations in thyroid hormone levels were actually due to NTIS, as opposed to an underlying previous clinical thyroid disease.^9^ So, these results have to be carefully analysed.

Our findings suggest that underlying hypothyroidism does not lead to worse outcomes in patients with COVID-19. In fact, those patients presented indicators of better outcomes than patients without hypothyroidism. At hospital admission, respiratory rates over 24 breaths per minute were less frequent in hypothyroidism patients than in the control group (36.1% vs 42.0%; p=0.050). It is known that tachypnea is one of the alarming COVID-19 symptoms as it may be an indicator of COVID-19 pneumonia.^39^ Despite the fact that SF ratio, a strong predictor of COVID-19 prognosis and mortality,^4,40^ was slightly higher in hypothyroidism patients, the difference was not clinically relevant (428.6 vs. 423.8, p=0.034).

The lower requirement of mechanical ventilation had not been observed previously, and it was quite unexpected. It is known that patients with decompensated hypothyroidism may have abnormalities in pulmonary function, muscle myopathy and neuropathy, with respiratory muscle weakness, but these abnormalities improve with treatment.^41,42^ In a previous propensity score matching analysis of patients from a New York health system, adjusted for age, sex, race, body mass index, smoking status, and number of comorbidities, hypothyroidism was not associated with an increased risk of mechanical ventilation (adjusted OR: 1.17 [95% CI: 0.81–1.69]) or death (adjusted OR: 1.07 [95% CI: 0.75–1.54]). However, this analysis was limited by the sample size of patients who were hospitalized, limited number of events, and fixed selection of three controls per case, which forces selection of controls.^11^ The other previous cohort studies and case series were also limited by small sample sizes and lack of controls.^8,9,11,13,31,43,44^ In the present study, a caliper width of 0.17 was used for the matched analysis. It is considered conservative and restricts the selection of controls which are indeed similar to the cases, as recommended.^45^ Due to a successful matched-analysis between samples, it is unlikely that age, sex or other underlying diseases have interfered with the results, although patients with hypothyroidism showed a higher prevalence for coronary artery disease and chronic kidney disease. Medications used during hospitalization were also similar in both groups, and, therefore, cannot explain the difference in prognosis as well.

Study limitations include its retrospective design, with data being collected by chart review only, and the fact that thyroid disease status was based on patients information given at hospital admission, so it was not possible to know if the hypothyroidism was compensated or not. Nevertheless, to the best of our knowledge, the study has the largest sample of COVID-19 patients with hypothyroidism published so far. TSH and thyroid hormone (T4 and T3) levels were not evaluated during the time of hospitalization and time in the ICU, as this is not recommended during hospitalization due to Covid-19 or other acute diseases is by the health organizations, and it was not our aim to access the effects of NTIS. The assessment of thyroid hormone levels throughout the course of Covid-19 is still challenging due to the possibility of overdiagnosis due to abnormal hormone levels during acute systemic disease. It is important to emphasize that these hormonal changes are adaptive processes and cannot be defined as a disease itself.

## CONCLUSION

In this multicenter cohort of Covid-19 in patients, those with underlying hypothyroidism, had similar comorbidities, clinical presentation and laboratory exams than the control group. It was found, nonetheless, that a lower proportion of patients with hypothyroidism required mechanical ventilation, and had a trend of lower in-hospital mortality. Therefore, hypothyroidism does not seem to be associated with a worse prognosis, and should not be considered among the comorbidities that indicate a risk factor for COVID-19 severity.

## Data Availability

Data is available upon reasonable request.

## DECLARATIONS

### Ethics approval

The study has been conducted by a protocol approved by the National Commission for Research Ethics^15^ (CAAE 30350820.5.1001.0008). Individual informed consent was waived due to the pandemic and to the fact that all data collected was unidentified, gathered through medical records.

### Availability of data and materials

Data is available upon reasonable request.

### Competing interests

The authors declared no potential conflicts of interest with respect to the research, authorship, and/or publication of this article.

### Funding

This study was supported in part by Minas Gerais State Agency for Research and Development (*Fundação de Amparo à Pesquisa do Estado de Minas Gerais - FAPEMIG*) [grant number APQ-00208-20], National Institute of Science and Technology for Health Technology Assessment (*Instituto de Avaliação de Tecnologias em Saúde – IATS*)/ National Council for Scientific and Technological Development (*Conselho Nacional de Desenvolvimento Científico e Tecnológico - CNPq*) [grant number 465518/2014-1 and 147122/2021-0].

DNP was funded by a research scholarship from IATS/CNPq [grant number 147122/2021-0].

### Role of the funder/sponsor

The sponsors had no role in study design; data collection, management, analysis, and interpretation; writing the manuscript; and deciding to submit it for publication. DNP, LFGS, MMMG, MSM and MCP had full access to all the data in the study and had responsibility for the decision to submit for publication.

### Authors contributions

Substantial contributions for the conception or design of the manuscript: MSM, DNP, LFGS and MMMG.

Substantial contributions for data acquisition, analysis or interpretation: all authors. Manuscript formulation: MSM, DNP, LFGS, MMMG and MCP.

Critically revised the manuscript in relation to the important intellectual content: all authors.

Final version’s approval: all authors

Agreed to be responsible for all aspects of the work, ensuring that issues related to precision of integrity in any of the work’s parts will be properly investigated and solved: MSM and MCP.

## Acknowledgments

We would like to thank the hospitals which are part of this collaboration, for supporting this project: Hospitais da Rede Mater Dei; Hospital das Clínicas da UFMG; Hospital de Clínicas de Porto Alegre; Hospital Santo Antônio; Hospital Eduardo de Menezes; Hospital Tacchini; Hospital Márcio Cunha; Hospital Metropolitano Dr. Célio de Castro; Hospital Metropolitano Odilon Behrens; Hospital Risoleta Tolentino Neves; Hospital Santa Rosália; Hospital Santa Cruz; Hospital São João de Deus; Hospital Semper; Hospital Unimed-BH; Hospital Universitário Canoas; Hospital Universitário Santa Maria; Hospital Moinhos de Vento; Hospital Luxemburgo; Instituto Mario Penna; Hospital Nossa Senhora da Conceição; Hospital João XXIII; Hospital Júlia Kubitschek; Hospital Mãe de Deus; Hospital Regional do Oeste; Hospital das Clínicas da Faculdade de Medicina de Botucatu;Hospital Universitário Ciências Médicas; Hospital Bruno Born; Hospital SOS Cárdio; Hospital São Lucas da PUCRS.

We also thank all the clinical staff at those hospitals, who cared for the patients, and all undergraduate students who helped with data collection.

## Transparency declaration

The lead authors (DNP, MCP and MSM) affirm that the manuscript is an honest, accurate, and transparent account of the study being reported; that no important aspects of the study have been omitted; and that any discrepancies from the study as originally planned (and, if relevant, registered) have been explained.

### LIST OF ABBREVIATIONS

COVID-19: Coronavirus disease 19
NTIS: Non-thyroidal illness syndrome
T3: Triiodothyronine
T4: Thyroxine
TSH: Thyroid-stimulating hormone
ICU: Intensive Care Unit
REDCap: Research Data Capture
COPD: Chronic obstructive pulmonary disease
SF ratio: Peripheral oxygen saturation/fraction of inspired oxygen ratio
GCS: Glasgow coma score
HR: Heart rate
RR: Respiratory rate
Sat O_2_: Peripheral capillary oxygen saturation
aPTT: Activated partial thromboplastin time
AST: Aspartate transaminase
ALT: Alanine aminotransferase
BNP: B-type natriuretic peptide
CPK: Creatine phosphokinase
NT-proBNP: N-terminal pro-brain natriuretic peptide
INR: International normalized ratio
VTE: Venous thromboembolism
ACE 2: Angiotensin-converting enzyme 2

## Supplementary

**Table S1-.** Medications used during hospitalization.

| Medications | Hypothyroidism<br>N = 526<sup>1</sup> | Control Patients<br>N = 526<sup>1</sup> | p-value<sup>2</sup> |
| --- | --- | --- | --- |
| **Medications used for COVID-19 management\*** |  |  |  |
| Azithromycin | 393 (74.7%) | 409 (77.8%) | >0.999 |
| Chloroquine | 6 (1.1%) | 7 (1.3%) | >0.999 |
| Hydroxychloroquine | 41 (7.8%) | 48 (9.1%) | 0.085 |
| Interferon | 2 (0.4%) | 0 (0.0%) | >0.999 |
| Convalescent plasma | 8 (1.5%) | 5 (1.0%) | >0.999 |
| Remdesivir | 0 (0.0%) | 1 (0.2%) | >0.999 |
| Ritonavir/Lopinavir | 1 (0.2%) | 1 (0.2%) | >0.999 |
| Clarithromycin | 19 (3.6%) | 24 (4.6%) | >0.999 |
| **Other medications used during hospitalization** |  |  |  |
| Antiarrhythmic | 50 (9.5%) | 44 (8.4%) | 0.089 |
| Antibiotics** | 445 (84.6%) | 463 (88.0%) | >0.999 |
| Anticoagulation | 469 (89.2%) | 483 (91.8%) | >0.999 |
| Prophylactic | 393 (74.7%) | 405 (77.0%) | >0.999 |
| Therapeutic | 79 (15.0%) | 88 (16.7%) | >0.999 |
| Antifungal | 24 (4.6%) | 31 (5.9%) | >0.999 |
| Inotropes | 128 (24.3%) | 157 (29.8%) | 0.044 |
| Non-steroidal anti-inflammatory | 87 (16.5%) | 65 (12.4%) | 0.144 |
| Corticoids*** | 379 (72.1%) | 363 (69.0%) | >0.999 |
| Dexamethasone | 326 (62.0%) | 310 (58.9%) | >0.999 |
| Other corticoids | 88 (16.7%) | 73 (13.9%) | >0.999 |
| Statin | 103 (19.6%) | 73 (13.9%) | 0.167 |

<sup>1</sup> n (%) <sup>2</sup>Wilcoxon rank sum test; Pearson's Chi-squared test; Fisher's exact test
\*Sarilumab, Favipiravir, Immunoglobulin, Tocilizumab and Umifenovir were not used by any patients.
\*\* Except for Azithromycin and Clarithromycin
\*\*\* Some patients used more than one type of corticoid

## Notes

### Competing Interest Statement

The authors have declared no competing interest.

### Funding Statement

This study was supported in part by Minas Gerais State Agency for Research and Development (Fundacao de Amparo a Pesquisa do Estado de Minas Gerais - FAPEMIG) [grant number APQ-00208-20], National Institute of Science and Technology for Health Technology Assessment (Instituto de Avaliacao de Tecnologias em Saude - IATS)/ National Council for Scientific and Technological Development (Conselho Nacional de Desenvolvimento Cientifico e Tecnologico - CNPq) [grant number 465518/2014-1 and 147122/2021-0].
DNP was funded by a research scholarship from IATS/CNPq [grant number 147122/2021-0].

### Author Declarations

The study has been conducted by a protocol approved by the National Commission for Research Ethics (CAAE 30350820.5.1001.0008). Individual informed consent was waived due to the pandemic and to the fact that all data collected was unidentified, gathered through medical records.

